# Superspreaders help covid-19 elimination

**DOI:** 10.1101/2020.04.19.20071761

**Authors:** David Kault

## Abstract

There seems to be widespread pessimism regarding the ability of a nation to eliminate covid. One factor in this pessimism seems to be concern that covid might always be able to re-emerge because of the ongoing presence of unrecognised asymptomatic cases. However, it is shown here that it should be possible to eliminate covid more easily than anticipated, for a reason that at first glance seems paradoxical - the presence of superspreaders. If superspreaders are responsible for most of the spread, then, with the average number of secondary cases fixed at say *R*_0_ = 2.5, we have to conclude that superspreaders are relatively rare. When towards the end of an elimination program, there are very few infected people, whether symptomatic or asymptomatic, that small number of people may well not include any superspreaders. As a result, chance effects may make extinction likely. Nevertheless it is clear an attempt at elimination will require a rather onerous “lockdown”. In this paper we use a branching processes model to look at the tradeoff between risk of disease re-emergence and the length of “lockdown” required after a program of elimination has dropped the number of symptomatic cases in a region to just one.

## Introduction

The experience of Taiwan, China, Vietnam, New Zealand and perhaps Australia indicates that covid-19 can be eliminated. Elimination may require lowering the reproductive rate of the infection by “lockdown” measures until there are very few active cases and then perhaps waiting for several incubation periods without new cases before ending the lockdown.

When numbers are large, each generation of cases will increase by a factor of ≈ *R*_0_ (ignoring saturation effects). However, when numbers are very low, stochastic effects are important, particularly if superspreaders are prominent. To take an extreme example, it is clear that if superspreaders were all important, so that the average *R*_0_ of say 2.5 was made up of 1% of infected people, who each spread the disease to 250 others and 99% who spread the disease to no-one, then there is at least a 99% chance that any randomly chosen infected person will not generate ongoing transmission. In general, a degree of superspreading, seems to be important across a range of infectious disease.^[1, 2]^ A negative binomial distribution seems to approximate the distribution of the number of secondary cases generated by a primary case of an infectious disease.^[1, 2]^ This distribution depends on the average number infected *R*_0_ and a dispersion parameter *r* (some authors use *k* in place of *r*). With *R*_0_ fixed, if *r* → ∞, there is no superspreading phenomenon and the number of secondary cases is the total randomness described by a Poisson distribution. If *r* → 0, superspreading becomes increasingly important, with a smaller and smaller proportion of infectious cases spreading the disease to more and more people. Likewise, as *r* → 0, if we start with a randomly chosen infected person, the chance that they are a superspreader becomes smaller and so the chance that they give rise to ongoing disease transmission reduces. Correspondingly, with very low numbers of infected people close to the end of a program of elimination, as *r* → 0, the probability of elimination becomes high.

There are various estimates of the dispersion parameter *r* for the distribution of secondary covid cases that have occurred from various primary cases. For the distribution before lockdown there is an estimate that *r* = 0.1 from observation of the initiation of the epidemic in various countries.^[3]^ A paper based on data from the early epidemic in China, states that there will be a probability of < 50% of covid elimination if there are at least 4 cases present.^[4]^ It is difficult to find the direct estimate of *r* in this paper but this probability of elimination is compatible with an *r* value of between 0.126 and 0.172. A paper based on data after lockdown in China states that 80% of cases are caused by 8.9% of infected people.^[5]^ With *R*_0_ also specified, it is not possible to precisely match the two figures 80% and 8.9% by varying the single parameter *r*, though *r* = 0.126 provides a close approximation. (Unfortunately, the paper directly states a negative binomial dispersion parameter value of 0.58, but this is not compatible with their data.) The parameter *r* in the case of SARS is estimated to be 0.16-0.17^[2]^ A million fold variation in viral load in the secretions of covid patients also suggests a prominent role for superspreaders.^[6]^ Given these various estimates of *r*, it seems that the true distribution of the number of secondary cases of covid generated by various primary cases, can be approximated by a negative binomial distribution with dispersion parameter *r* in the range 0.10 to 0.17. Accordingly, two negative binomial distributions, one with *r* = 0.10 and the other with *r* = 0.17, are used in a model of the last stage of a program of regional covid elimination.

For both distributions, the case of a policy of release of lockdown once there is just a single newly diagnosed symptomatic case, is examined. Results are obtained by analytical methods. The effect of two further policies are obtained by simulation. These policies are waiting one incubation period after this time before lifting restrictions and waiting two incubation periods before lifting restrictions.

This paper assumes that an attempt has been made to eliminate covid-19 from a region/province/nation. It assumes no appreciable herd immunity. It also assumes that there is no quarantine free cross border human travel into the region considered and this restriction will be maintained. We make the conservative assumption that the moment lockdown is lifted, transmission immediately goes back to its pattern at the beginning of the epidemic, which for both distributions is assumed to have *R*_0_ = 2.5.

It is known that a proportion of cases are asymptomatic. Here this proportion *p_a_*, is taken to be 50%. It is further assumed that the symptomatic cases up to their time of isolation and the asymptomatic cases, both have the same distribution for the number of secondary infections they generate. This too seems likely to be a conservative assumption.

## Methods

It is known from the theory of branching processes in discrete time, that the probability of extinction of a branching process is given by the (smallest) solution to the equation *ϕ*(*u*) = *u* where *ϕ*(*x*) is the probability generating function for the number of secondary cases of a single index case.^[7]^ This equation can be solved by Newton’s method. If the distribution of secondary cases is negative binomial with *R*_0_ = 2.5 and *r* = 0.10, the probability of extinction of a single branching process is 0.861. With the same *R*_0_ and *r* = 0.17 the probability of extinction is 0.784. We denote these probabilities of extinction of a line from a single case as *p_e_*.

However, once a closed region is down to a single newly diagnosed symptomatic case, we are assuming that there will be on average one unnoticed asymptomatic infectious person (*p_a_* = 50%). It is assumed, as an uninformative Bayesian prior, that the actual number of asymptomatic cases will follow a geometric distribution. Nevertheless, as described above, there may be no ongoing transmission from any one case and so it is possible that there will be no ongoing transmission from however many infectious people there are left. We make two conservative assumptions here. First, that the last symptomatic case has already definitely seeded one other case before diagnosis and isolation. Second, that with lockdown lifted as soon as the number of newly diagnosed symptomatic cases is down to 1, *R*_0_ for all infectious people still present, immediately goes back to 2.5. It is shown in Attachment 1 that the probability of epidemic re-emergence will then be 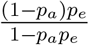. We denote this probability as *p_T_*_0_ where “*T*” denotes Total disease elimination and the “0” stands for release of lockdown zero incubation periods after the the number of newly diagnosed symptomatic cases has dropped to one.

There are two further policy scenarios considered - continuing restrictions for 1 incubation period after the last symptomatic person has been isolated, and continuing retrictions for 2 incubation periods. During this period of ongoing lockdown it is assumed that a negative binomial distribution of secondary cases will still apply but with *R*_0_ = 0.8. However, the dispersion parameter *r* is assumed to be unchanged. The probability that the disease will truly have been eliminated if no further symptomatic case emerges during this extended lockdown is calculated by simulation. The probability of total elimination of the disease after 1 and after 2 incubation periods is denoted *p_T_*_1_ and *p_T_*_2_ respectively. Each simulation employs 10 million runs. A fortran program corresponding to these simulations is given as an attachment.

## Results

Table: Results with *p_e_* and *p_T_*_0_ calculated analytically and *p_T_*_1_ & *p_T_*_2_ calculated by simulation. The analytic calculation assumes that the last diagnosed case has already seeded exactly one other case before isolation and then *R*_0_ immediately returns to 2.5. The simulations assume that during isolation, all infected people seed cases according to a distribution with *R*_0_ = 0.8, but afterwards *R*_0_ returns immediately to 2.5

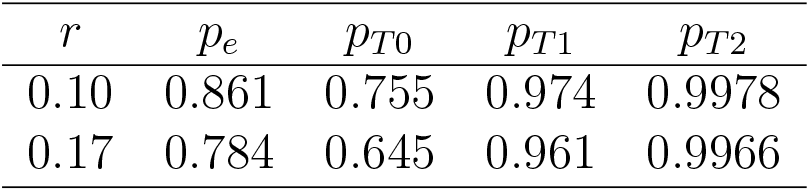

Unfortunately, in implementing policies whose outcomes are denoted *p_T_*_1_ and *p_T_*_2_, there will be occasions where relief from “lockdown” will be further delayed by re-emergence of symptomatic cases before the one or two incubation periods are completed. In the case of both the distributions, this happens about 31% and 34% of the time with a policy of a 1 or 2 incubation wait respectively.

## Discussion

A policy of elimination requires a more intense “lockdown” than a policy of “flattening the curve” so that the epidemic peak is within the capacity of a region’s ICU beds. However, a study by the Imperial College,^[8]^ shows that there are two rather fine lines between flattening the curve sufficiently and on the one hand, elimination, or on the other hand, an insufficiently flattened epidemic and a huge death rate. Even if “optimal” curve flattening can be achieved there will still be many deaths. In addition, whilst slightly less intense lockdown is required for curve flattening rather than elimination, the period of “lockdown” for curve flattening, will have to be continued for far longer. An advantage of the curve flattening approach is that curve flattening could lead to herd immunity and thus allow less restricted international travel to parts of the world that have not achieved elimination, though this prospect is uncertain. On the other hand, international travel without quarantine will not be possible between a country that has achieved elimination and a country which has not, at least until a vaccine is available. For this author, the arguments in favour of a policy of elimination are ovewhelming, for nations/regions which have the resources to implement it and who have secure borders. New Zealand is following a policy of elimination and there seems to be emerging realisation that this is a real option for Australia. Since the intensity of “lockdown” required for elimination will be quite onerous, it is important that the length of “lockdown” required is balanced against the probability that this lockdown period will be successful in eliminating the virus.

The calculations show that there is better than 99.7% chance of elimination if two incubation periods pass without re-emergence of symptomatic cases. It would seem that, when numbers of symptomatic cases have been reduced to rather low levels, lockdown periods, could be decreased further by subdividing a region. A region implementing an elimination strategy could be subdivided into subregions which could be temporarily disconnected from each other in terms of human travel, such that each subregion has only one case. Relief from severe restrictions could then commence quite quickly.

There are some clear limitations to this study. No sensitivity analysis has been done beyond calculations for the two specific distributions described. In particular, there is no sensitivity analysis for likely different values for the proportion asymptomatic *p_a_*. However, the fortran programs below could facilitate such a sensitivity analysis if there was more knowledge about the range of plausible values.

The calculations and simulations also assume discrete time steps and a different analysis will be required to account for a more realistic model in continuous time.^[9]^ However, one might reasonably hope that more realistic simulations in continuous time will yield fairly similar results to these simpler simulations - but this would need to be checked.

There are some assumptions that are too pessimistic. Some assumptions that are likely too conservative have been mentioned above, but there are others. For example, many asymptomatic cases may be detected by contact tracing. There is also implicitly some assumptions that may be too optimistic. For example, it is assumed that there are not cases of prolonged infectious periods or human carrier states or animal reservoirs of the virus in the region. Nevertheless this study gives cause for optimism about the length of lockdown required for any region attempting to eliminate this disease.

## Data Availability

There is no relevant data as such - just computer programs developed for this project. The main programs are included as an attachment at the end of the paper. Programs which are near duplicates of each other or that are almost trivial are omitted, but the whole suit of programs used here are available from the author.

## Acknowledgements

I thank Profs Shijit Kumar and James McCaw of Melbourne University for their helpful comments.

## Attachment 1

It is assumed that the number of asymptomatic cases is given by a geometric distribution, so that the probability that there are k asymptomatic cases is 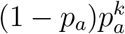, *k* = 0, 1, 2,….

The probability *p_T_*_0_, that none of these cases gives rise to ongoing transmission is found by summing the probability over all possible values of *k* that there are k such cases and that transmission from all such cases goes extinct with probability 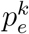 this gives 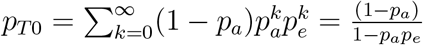

If we include the conservative assumption that before diagnosis, the last case diagnosed had definitely seeded a single case, we have the result 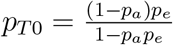

## Attachment 2

Fortran program using Newton’s method to calculate the probability of extinction of the branching process for various probability distributions

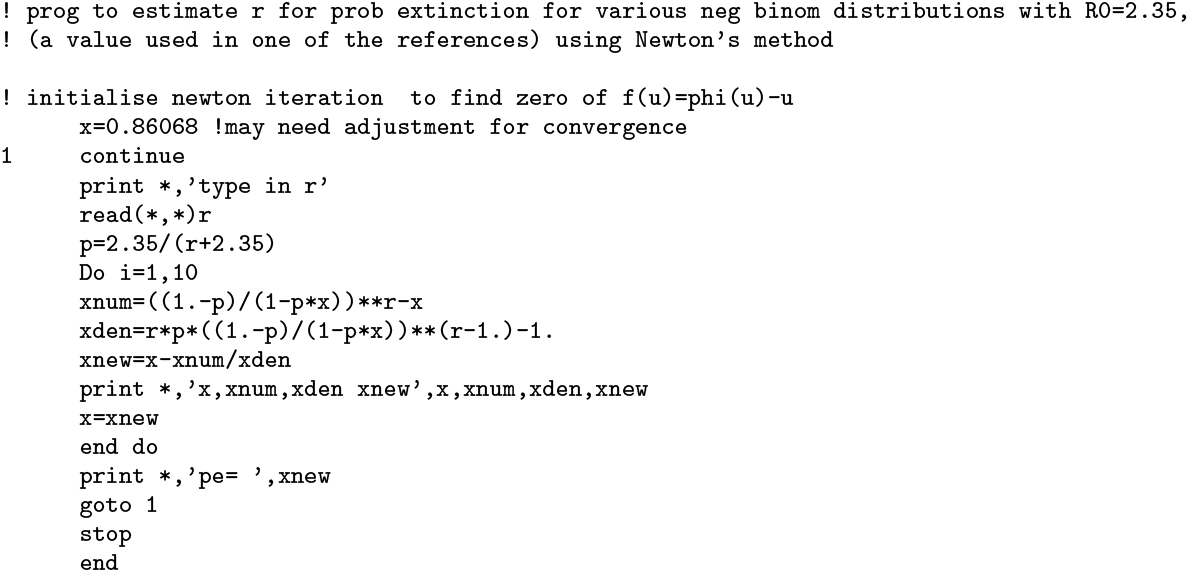

The fortran program below calculates by simulation, the probability of elimination when there are two incubation periods in which, under “lockdown” conditions, number of secondary cases are determined by a negative binomial distribution, with *R*_0_ reduced from 2.5 to 0.8, but with *r* unchanged unchanged. Since this negative binomial is used repeatedly in the simulation, it is useful to calculate its cumulative distribution using a separate program. A few other very short, almost trivial, programs are also used in this project. They are not copied here but are available on request to the author.

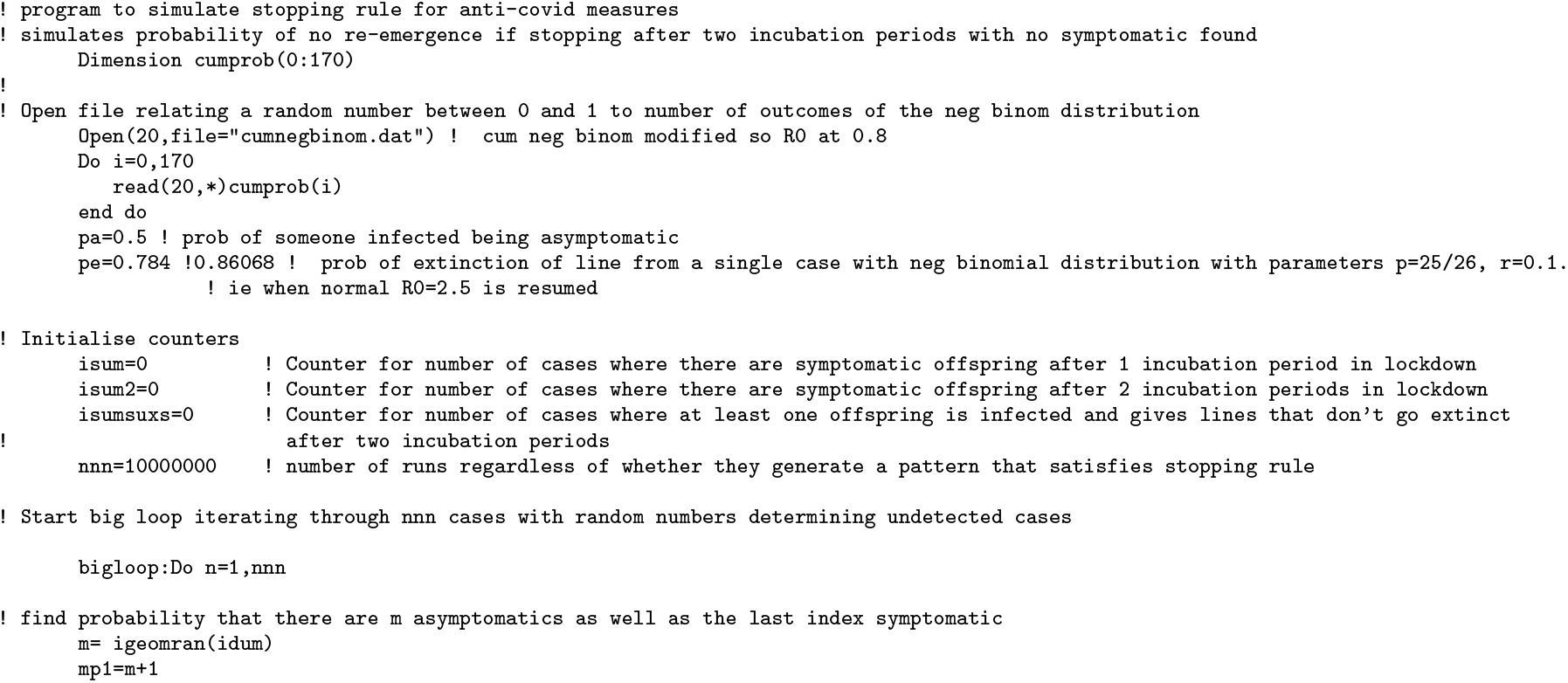

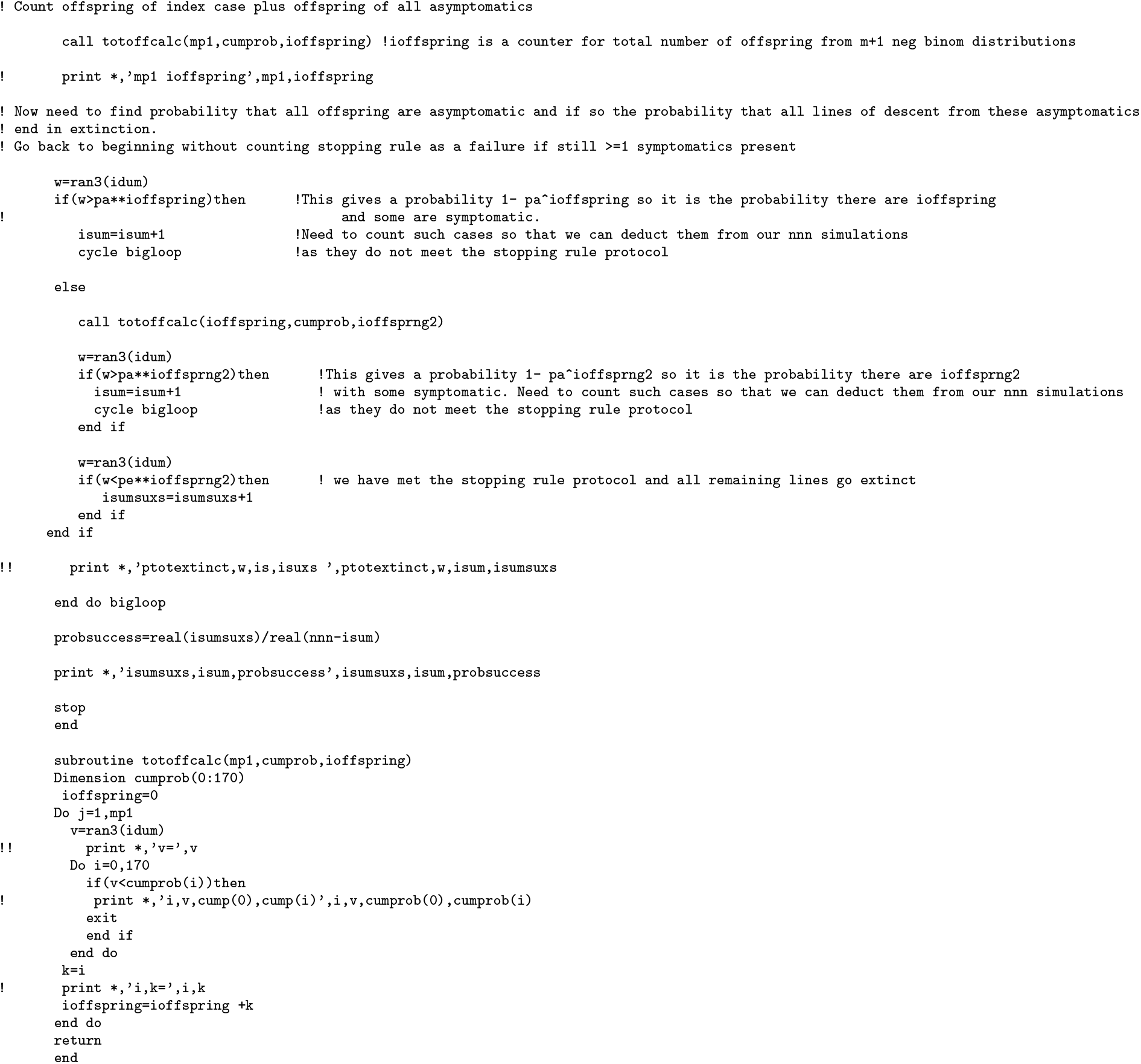

## Notes

### Competing Interest Statement

The authors have declared no competing interest.

### Funding Statement

Paper was prepared as part of my role as an adjunct senior lecturer, with no funding other than being allowed ongoing access to university library resources

